# Prevalence and risks of severe events for cancer patients with COVID-19 infection: a systematic review and meta-analysis

**DOI:** 10.1101/2020.06.23.20136200

**Authors:** Qiang Su, Jie-xuan Hu, Hai-shan Lin, Zheng Zhang, Emily C. Zhu, Chen-guang Zhang, Di-ya Wang, Zu-hua Gao, Bang-wei Cao

**Author notes:** Correspondence authors E-mail address for Dr Chenguang Zhang; for Dr Di-ya Wang; for Dr Zu-hua Gao; for Dr Bang-wei Cao.

## Abstract

**Background:** The corona virus disease 2019 (COVID-19) pandemic poses a severe challenge to public health, especially to those patients with underlying diseases. In this meta-analysis, we studied the prevalence of cancer among patients with COVID-19 infection and their risks of severe events.

**Methods:** We searched the Pubmed, Embase and MedRxiv databases for studies between December 2019 and May 3, 2020 using the following key words and terms: sars-cov-2, covid-19, 2019-ncov, 2019 novel coronavirus, corona virus disease-2019, clinical, clinical characteristics, clinical course, epidemiologic features, epidemiology, and epidemiological characteristics. We extracted data following PICO (patient, intervention, comparison and outcome) chart. Statistical analyses were performed with R Studio (version 3.5.1) on the group-level data. We assessed the studies’ risk of bias in accordance to the adjusted Joanna Briggs Institute. We estimated the prevalence or risks for severe events including admission into intensive care unit or death using meta-analysis with random effects.

**Findings:** Out of the 2,551 studies identified, 32 studies comprising 21,248 participants have confirmed COVID-19. The total prevalence of cancer in COVID-19 patients was 3.97% (95% CI, 3.08% to 5.12%), higher than that of the total cancer rate (0.29%) in China. Stratification analysis showed that the overall cancer prevalence of COVID-19 patients in China was 2.59% (95% CI, 1.72% to 3.90%), and the prevalence reached 3.79% in Wuhan (95% CI, 2.51% to 5.70%) and 2.31% (95% CI, 1.16% to 4.57%) in other areas outside Wuhan in China. The incidence of ICU admission in cancer patients with COVID-19 was 26.80% (95% CI, 21.65% to 32.67%) and the mortality was 24.32% (95% CI, 13.95% to 38.91%), much higher than the overall rates of COVID-19 patients in China. The fatality in COVID patients with cancer was lower than those with cardiovascular disease (OR 0.49; 95% CI, 0.34 to 0.71; p=0.39), but comparable with other comorbidities such as diabetes (OR 1.32; 95% CI, 0.42 to 4.11; p=0.19), hypertension (OR 1.27; 95% CI, 0.35 to 4.62; p=0.13), and respiratory diseases (OR 0.79; 95% CI, 0.47 to 1.33; p=0.45).

**Interpretation:** This comprehensive meta-analysis on the largest number of patients to date provides solid evidence that COVID-19 infection significantly and negatively affected the disease course and prognosis of cancer patients. Awareness of this could help guide clinicians and health policy makers in combating cancer in the context of COVID-19 pandemic.

**Funding:** Beijing Natural Science Foundation Program and Scientific Research Key Program of Beijing Municipal Commission of Education (KZ202010025047).

## Introduction

The pandemic of severe acute respiratory syndrome corona virus 2 (SARS-CoV-2) has caused 4,794,932 confirmed cases and 316,420 deaths globally by May 7, 2020, affecting almost all the countries in the world.^1^ WHO announced COVID-19 as a public health emergency of international concern on January 30, 2020. Most COVID-19 cases experience mild to moderate respiratory symptoms or signs such as fever and cough, and will recover without special treatment. Unfortunately, the elder people, and those suffering underlying medical conditions such as cardiovascular disease, diabetes, chronic respiratory disease, and cancer are more likely to develop into more serious conditions, including multiple organ dysfunction syndrome or even death.^2-4^

Cancer is a major public health problem worldwide and is the second leading cause of death in the United States. Recent research had shown that there were about 24.5 million cancer cases worldwide and 9.6 million cancer deaths in 2017. In 2020, the number of new cancer cases and cancer related death is estimated to reach1,806,590 and 606,520 in the US, respectively.^5^Cancer patients are more susceptible to infections than those without cancer due to the systemic immunosuppression caused by the malignancy itself or anticancer treatments, such as chemo-radiation. ^6-9^ Therefore, it is conceivable that cancer patients are at higher risk of COVID-19 infection. Moreover, unplanned interruptions of regular anti-tumor therapies, due to the overloaded healthcare systems in the pandemic battle, expose cancer patients’ adverse clinical risks of uncontrolled primary disease. Furthermore, some tumor markers such as CEA, CA-125 can be elevated and positively correlated with the pathological progression of COVID-19 infection, which may increase the cancer detection rate. ^10^

The relationship between cancer and COVID-19 has drawn great attention from the beginning of the pandemic. An early study by Liang et al reported a cancer prevalence of 1.13% (95% CI, 0.61% to 1.65%) among 1,590 cases of COVID-19 in China (only 18 patients with cancer), which was higher compared with the overall cancer incidence of 0.29% in the Chinese population.^11^ More importantly, cancer patients with COVID-19 had a higher risk of severe events (a composite end point defined as the percentage of patients who were admitted to the intensive care unit (ICU) and required invasive ventilation or who died) compared with those COVID-19 patients without cancer (39% v 8%, HR: 5.34; 95%CI, 1.80 to 16.18; P = 0.0026). A pooled meta-analysis by Aakash Desai indicated that prevalence of cancer in COVID-19 patients was 2.0% (95% CI, 2.0% to 3.0%; I2 = 83.2%), while the prevalence was 3.0% (95% CI, 1.0% to 6.0%) in studies with a sample size smaller than 100, and 2.0% (95% CI, 1.0% to 3.0%) in studies with a sample size not smaller than 100.^12^ To guide clinical management and appropriate medical resources allocation, it is necessary to validate the relationship of cancer and COVID-19 in a much larger international scale. In the current study, we performed a meta-analysis of 21,248 participants in 32 clinical studies across six countries (China, Japan, Korea, Italy, France, USA)^11,13-43^to exploit the prevalence of cancer in COVID-19 patients as well as risks of severe events including ICU admission and death of these patients.

## Methods

### Search Methods and Study Selection

We searched Pubmed, Embase and MedRxiv databases for studies according to the following key words and terms: sars-cov-2, covid-19, 2019-ncov, 2019 novel coronavirus, Corona Virus Disease-2019, clinical, clinical characteristics, clinical course, epidemiologic features, epidemiology, epidemiological characteristics. The following data from all relevant studies between December 2019 and May, 2020 were included in our meta-analysis: (1) the study subjects were patients with confirmed diagnosis of SARS-CoV-2 viral infection, based on the detection of the viral nucleic acid by reverse transcription quantitative PCR (RT-qPCR) assay; (2) the results included age, sex, epidemiology and cancer information; (3) study types include cohort studies and clinical trials; (4) restriction of language to English only. Animal studies, systematic reviews, case reports, and) small studies of fewer than 10 patients were excluded. This study was performed according to the Preferred Reporting Items for Systematic Reviews and Meta-analyses (PRISMA) guideline.^44^ (eTable 1)

### Data extraction

The two authors (Q.S and JX.H) independently extracted the data by checking the titles, abstracts and full text. We then assessed the eligibility of the extracted studies using the PICO (patient, intervention, comparison and outcome) chart. Disagreements in the assessment were resolved via discussion with the third reviewer (CG.Z).^45^ Finally, they extracted the following information from all eligible studies: the first author, study period, district, gender, age, diagnosis methods, the number of total patients, the number of cancer patients, serious events, death toll, the number of comorbidities, and death toll of comorbidities.

### Data analysis

Statistical analyses were performed using R’s META package in R Studio (version 3.5.1). We focused on the following parameters in COVID-19 patients: the cancer prevalence, the ICU admission rate, and the cancer mortality. Different subgroups of patient population with cancer were compared. The prevalence for cancer in COVID-19 patients was expressed as percentage with 95% confidence interval (95% Cis). Odds ratio (OR) was used to estimate the -risks of ICU admission and death of cancer patients with COVID-19 compared with those suffering other comorbidities including cardiovascular diseases, diabetes, hypertension or non-cancer patients with COVID-19. Heterogeneity was evaluated using the Q test and Higgin’s and Thompson’s I2 (I2) statistics. Defined as the percentage of variations in the effect sizes that is not the cause of sampling errors, I2 statistics is robust and not sensitive to the number of studies under analysis. We consider I2 value of over 50% as moderate to high heterogeneity. P<0.05 was considered statistically significant. We used random-effects model if the I^2^ value was more than 50%. We also performed a multi-group subgroup analysis to explore the source of heterogeneity. Specifically, subgroup design was based on the sample size (smaller or greater than 100), and the area of the patients (China/Outside China; Japan/Korea/Italy/France/USA). The quality of the included studies was assessed by the Joanna Briggs Institute (JBI),^46^and we slightly modified the table to make it adapt with selected studies (eFigure 1). We also conducted sensitivity analysis by sequentially removing individual studies one by one, and evaluating the stability of the results. Begger test and Egger test were used to evaluate publication bias, caused by possibly missing unpublished studies of low effect sizes. Finally, we developed a funnel plot to describe the results of publication bias.

## Results

### Eligible Studies and Characteristics

Using our searching principles, we retrieved 2,551 studies from three databases between Dec 1st, 2019 and May 3rd, 2020, among which, 32 studies comprising 21,248 patients met our selection criteria (eTable 2). 22 of the 32 studies were mutually exclusive cohorts of patients in China (Wuhan n=13; outside of Wuhan n=5; unclear n=4), 2 studies based on cohorts of other Asian countries (Korean and Japan), and 8 studies were based in Europe and USA (Italy n=1; France n=1; USA n=6). The detailed research flow chart is shown in Figure 5, characteristics of the included studies are shown in Table 1, and data on severe events in patients with COVID-19 is shown in eTable3. Among the 21,248 patients in those 32 studies, valid data including basic information, epidemiological characteristics, clinical characteristics, prevalence and mortality of cancer or other complications were extracted from COVID-19 patients. All patients presented exhibited mild to severe clinical manifestations and were confirmed of COVID-19 infection RT-qPCR.

**Table 1.**
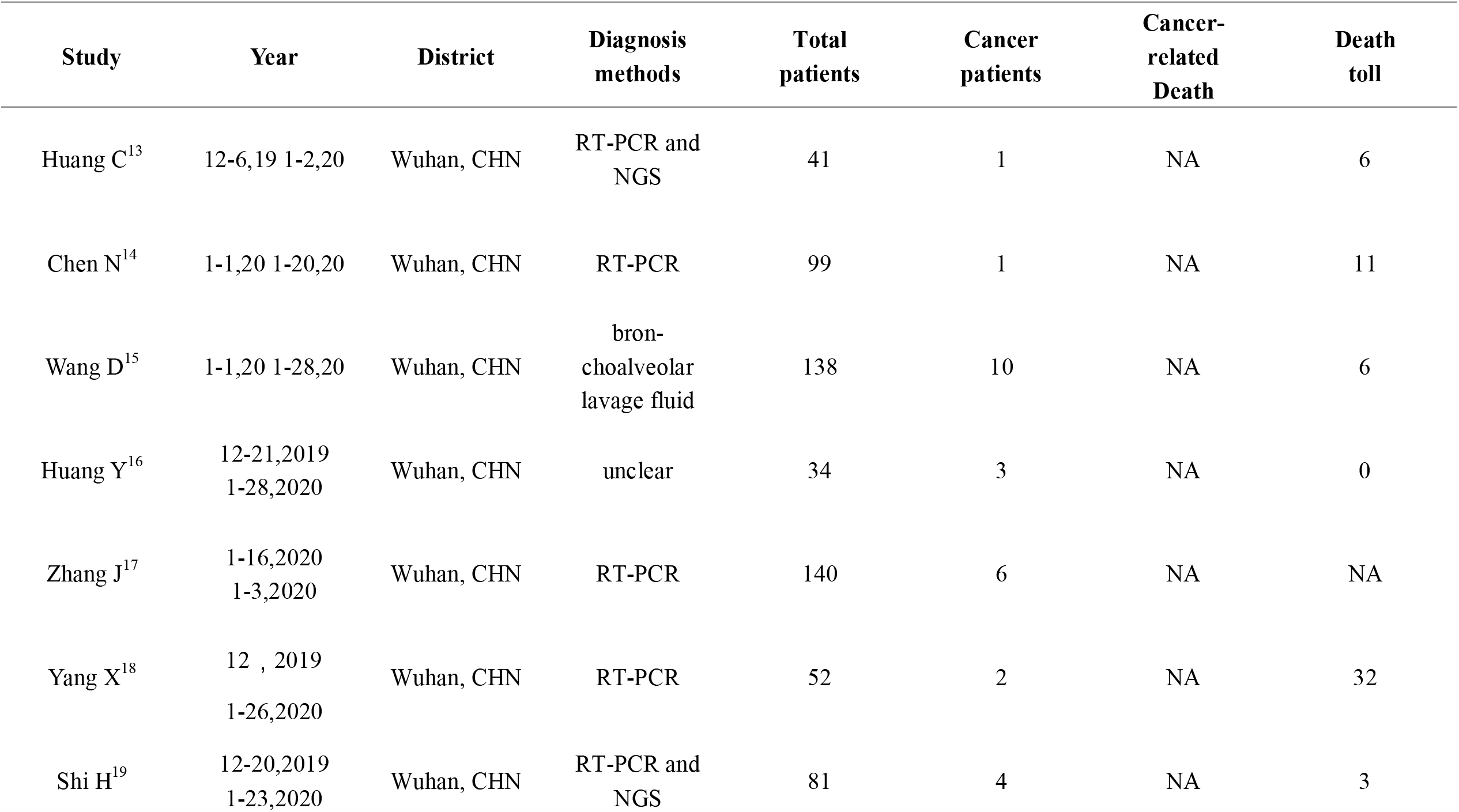
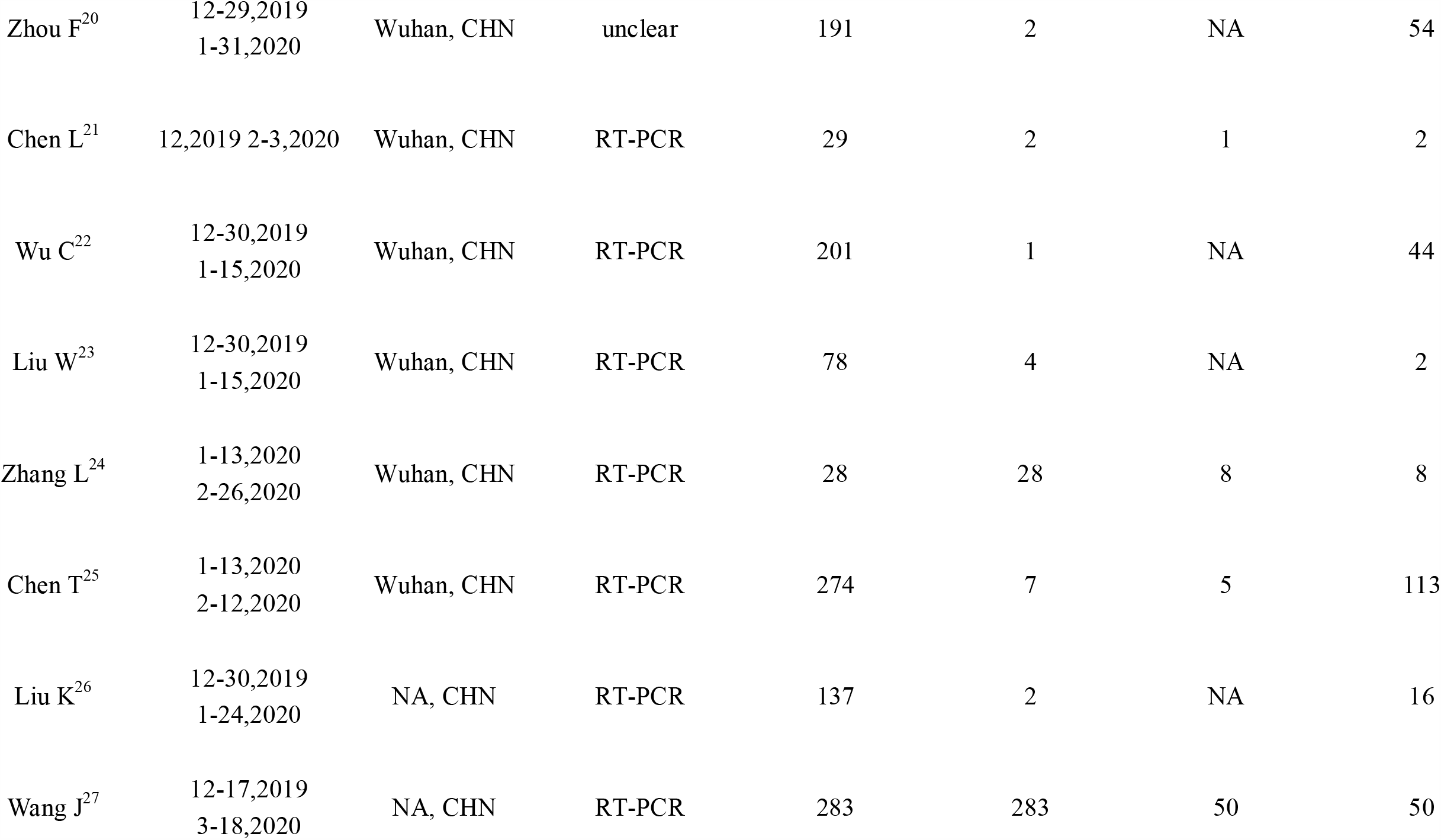
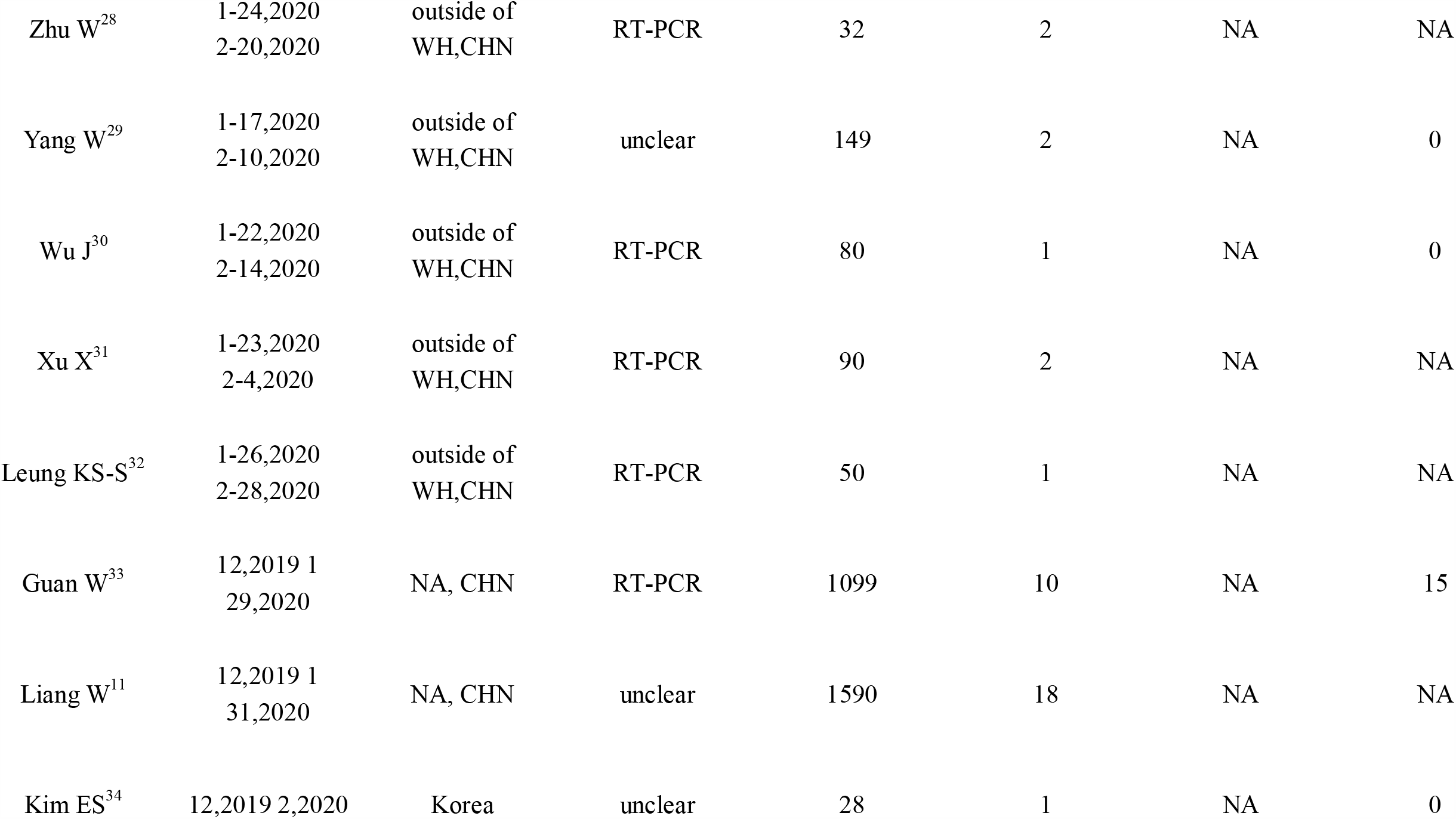
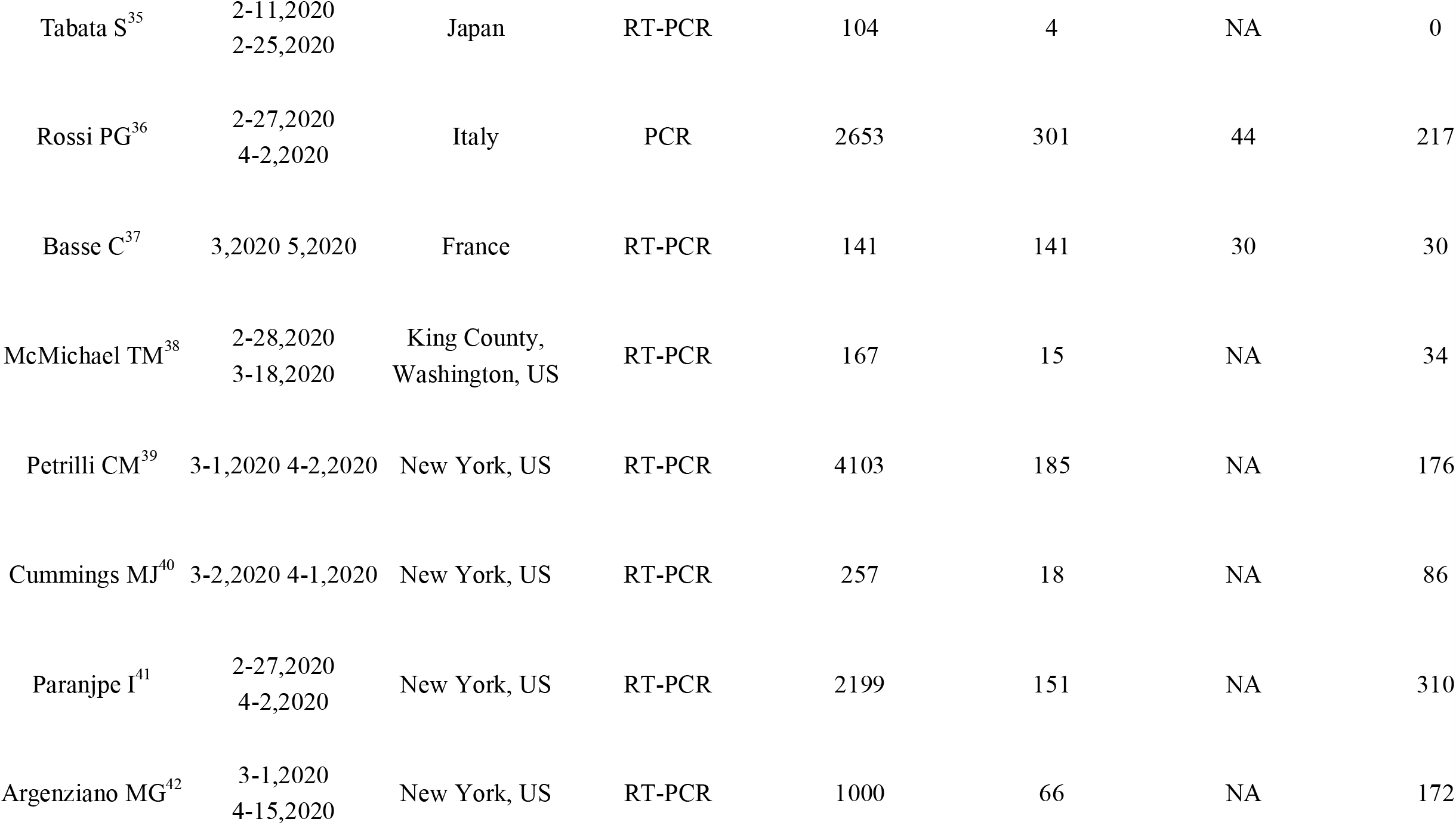
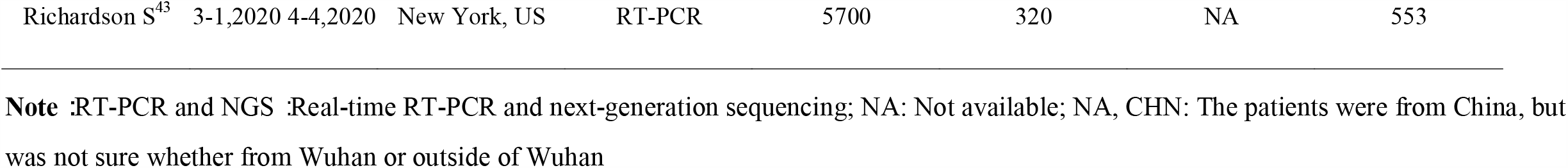
List of the 32 Studies Included in This Meta-Analysis.

**Figure 1.**
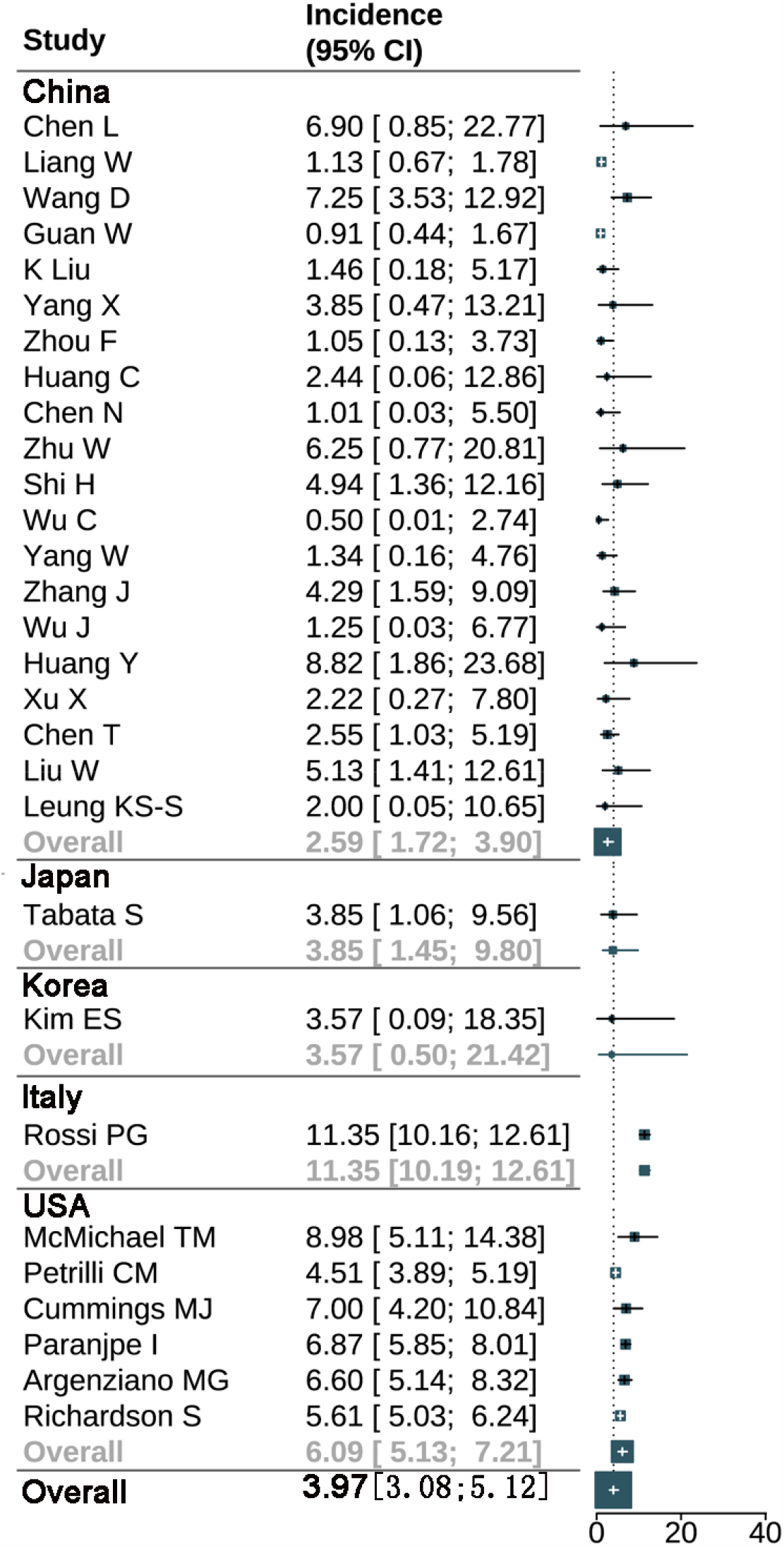
Prevalence of cancer in COVID-19 patients in China and outside China.

**Figure 2.**
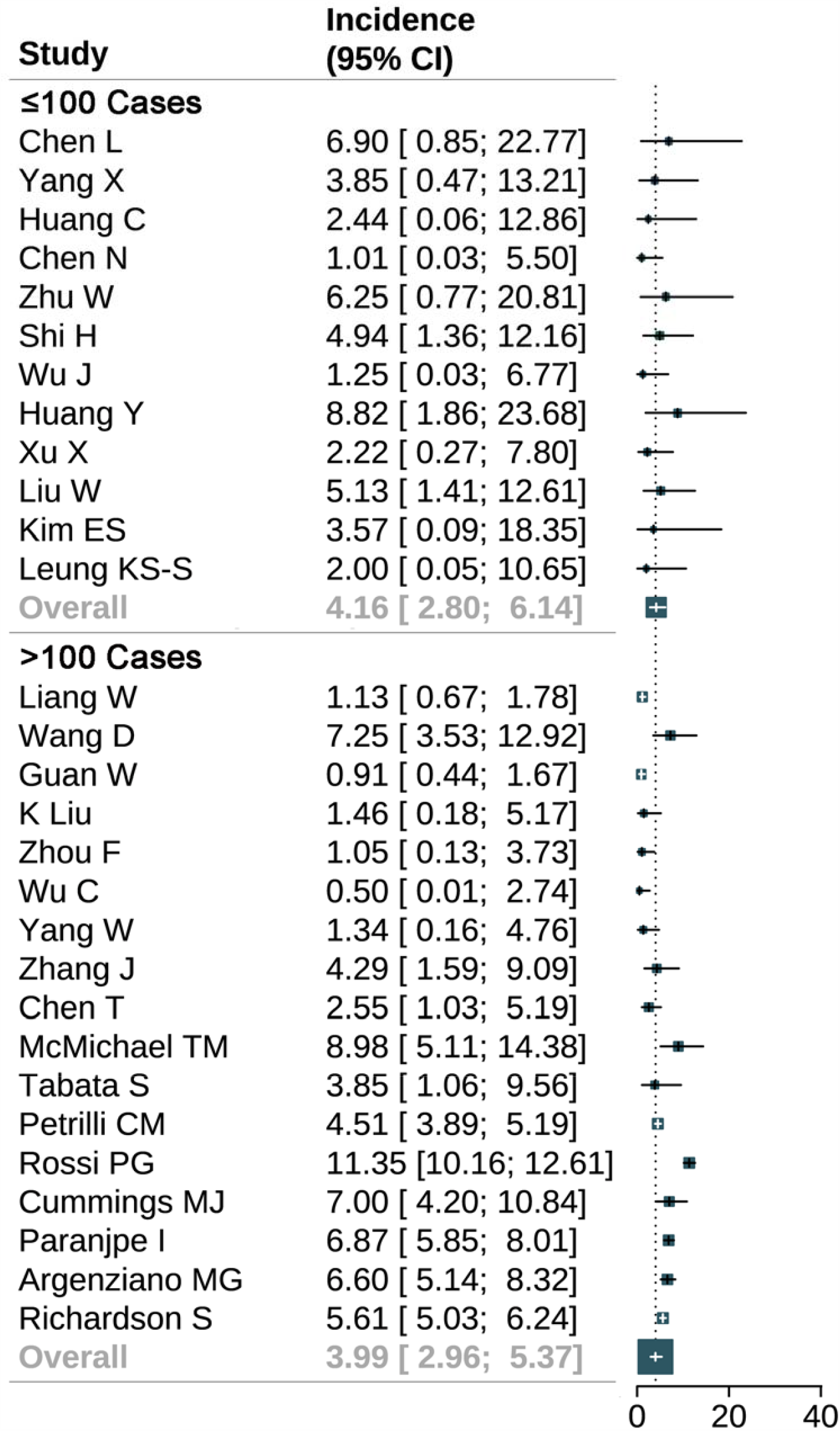
Prevalence of cancer in COVID-19 patients in subgroups based on sample size over or not over 100.

**Figure 3.**
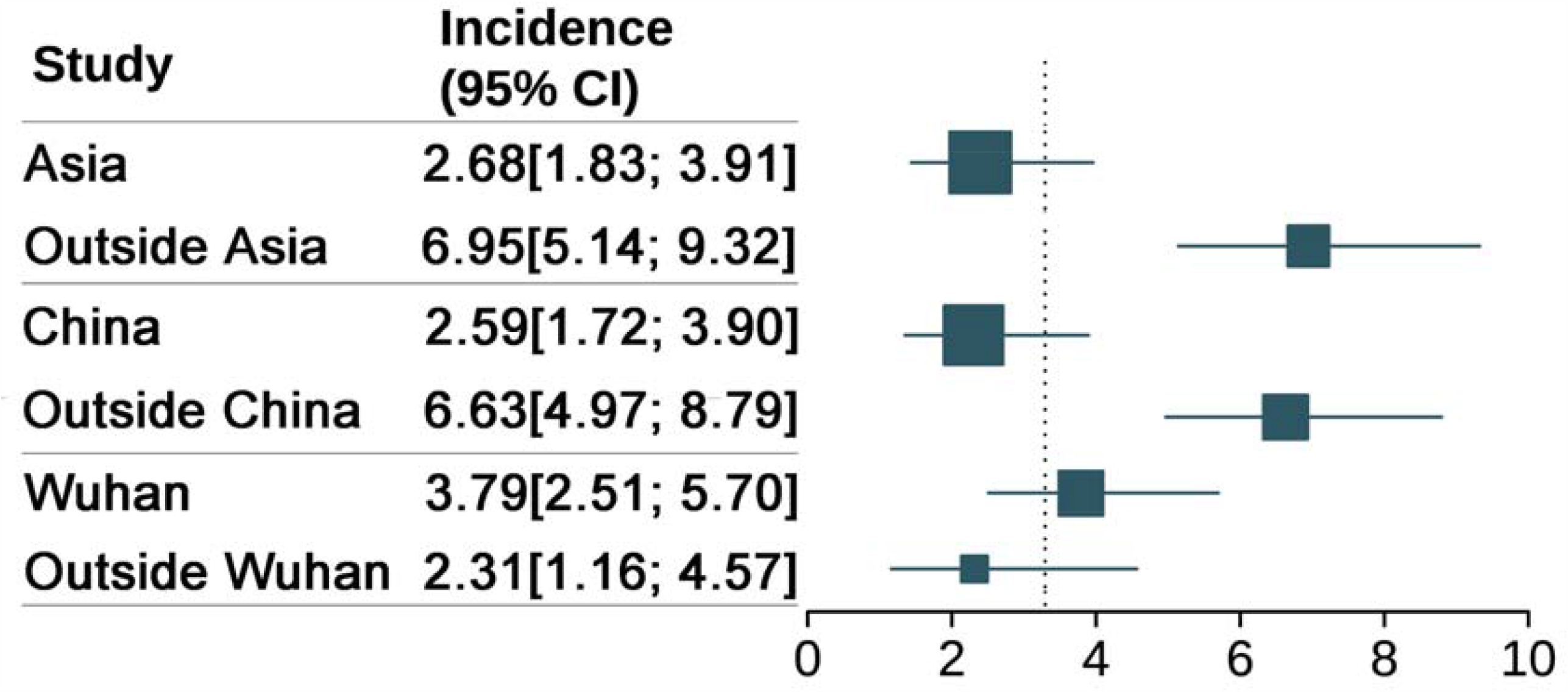
Prevalence of cancer in COVID-19 patients based on areas.

**Figure 4.**
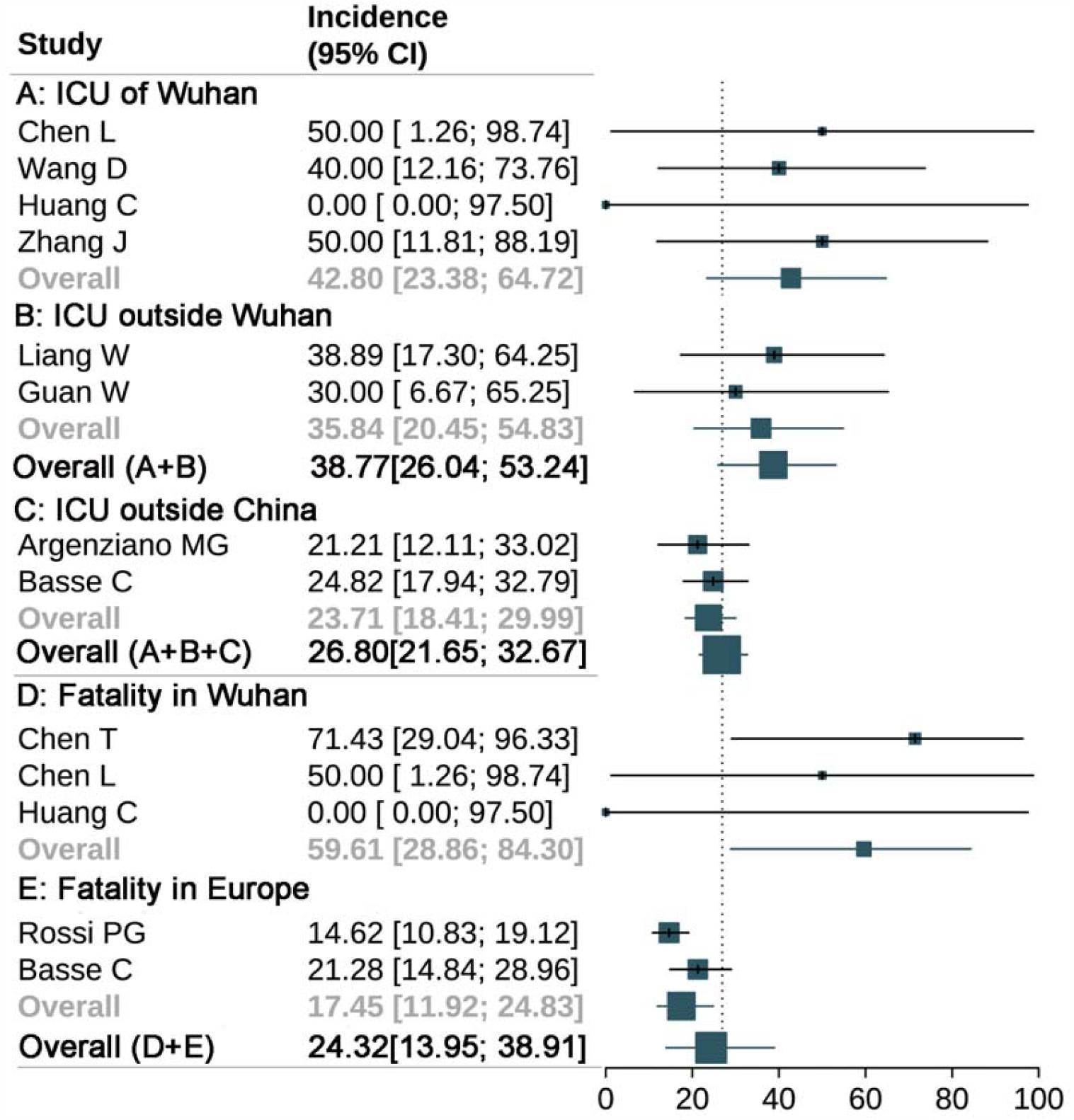
The incidence of ICU admission and fatality of cancer patients with COVID-19 in different areas. **A**:ICU admission incidence of cancer patients with COVID-19 in Wuhan, **B**:ICU admission incidence of cancer patients with COVID-19 outside Wuhan, **C**:ICU admission incidence of cancer patients with COVID-19 outside China, **A+B**:ICU admission incidence of cancer patients with COVID-19 in China, **A+B+C**:ICU admission incidence of cancer patients with COVID-19 in all areas(all available statistics), **D**:Fatality of cancer patients with COVID-19 in Wuhan; **E**:Fatality of cancer patients with COVID-19 in Europe; **D+E**: Fatality of cancer patients with COVID-19 in all areas(all available statistics)

**Figure 5.**
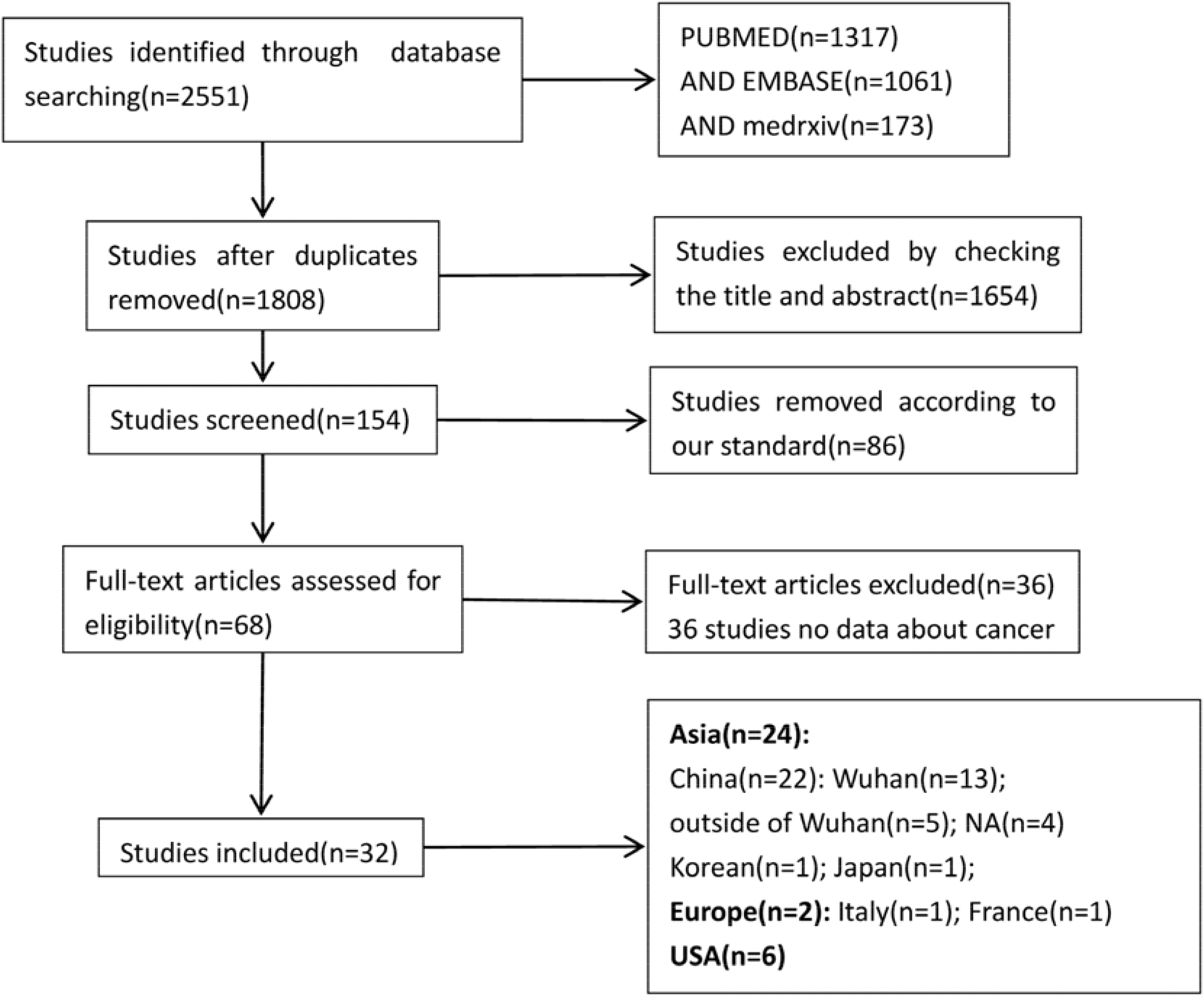
Flowchart depicting the studies selection process.

### Result of meta-analyses

#### Prevalence of cancer in COVID-19 patients

We first analyzed the cancer prevalence rates in COVID-19 patients. As shown in Figure 1, the total prevalence of cancer in COVID-19 patients was 3.97% (95% CI, 3.08% to 5.12%), which is higher than the total cancer rate (0.29%) in China.

When analyzing in subgroups of different sample size, the prevalence showed some variation. The prevalence of cancer was 4.16% (95% CI, 2.80% to 6.14%) in the subgroup whose sample size was ≤100, and 3.99% (95% CI, 2.96% to 5.37%) in the subgroup whose sample size was >100 (Figure 2). This observation is consistent and comparable with the analysis by Aakash Desai.^12^

We then analyzed the cancer prevalence based on patients’ geographic location. The overall cancer prevalence of COVID-19 patients was 2.59% (95% CI, 1.72% to 3.90%) in China, 3.85% (95% CI, 1.45% to 9.80%) in Japan, and 3.57% (95% CI, 0.50% to 21.42%) in Korea (Figure 1). The prevalence was higher in Italy and USA, which reached 11.35% (95% CI, 10.19% to 12.61%) and 6.09% (95% CI, 5.13% to 7.21%), respectively (Figure 1). Moreover, the cancer prevalence of COVID-19 patients in Wuhan was 3.79% (95% CI, 2.51% to 5.70%), higher than the average level of China (Figure 3).

Our analyses clearly showed that the cancer prevalence of COVID-19 patients was higher than that of the general population. In USA and European countries, the prevalence was higher than in China and other Asian countries. In China, the prevalence in Wuhan was higher than in other areas of China (Figure1 and 3).

#### Risk of severe events of cancer in COVID-19 patients

In China, the incidence of ICU admission in COVID19 patients with cancer was 38.77% (95% CI, 26.04% to 53.24%) (Figure 4). In Wuhan, the incidence of ICU admission and death in COVID19 patients with cancer were 42.80 (95% CI, 23.38 % to 64.72%) and 59.61% (95% CI, 28.86% to 84.30%) respectively. The incidence of ICU admission in COVID19 patients with cancer were 23.71% (95% CI, 18.41% to 29.99%) outside China, and the fatality reached 17.45% (95% CI, 11.92% to 24.83%) in Europe. The COVID19 patients in Wuhan apparently had a worse outcome than the rest of the China. Moreover, COVID-19 patients with cancer had a significantly higher ICU admission rate than those without cancer in China (OR 2.77; 95% CI, 1.28 to 6.00; p=0.46), and had overall higher fatality irrelative with areas (OR 4.87; 95% CI, 1.52 to 15.59; p=0.74/OR 3.97; 95% CI, 1.10 to 14.32; p=0.88) (eFigure 4). This may imply the existence of complex interrelationship between COVID-19 and cancer, where cancer was able to exaggerate COVID-19 infection and vice versa.

We further compared the rate of ICU admission rate and death of cancer with other underlying diseases in COVID-19 patients. As shown in eFigure 2, there was no significantly difference in the ICU admission rate of COVID 19 patients with cancer from COVID 19 patients with other comorbidities, including cardiovascular diseases (OR 0.89; 95% CI, 0.52 to 1.53; p=0.91), diabetes (OR 0.81; 95% CI, 0.48 to 1.37; p=0.85), hypertension (OR 0.88; 95% CI, 0.53 to 1.44; p=0.64), and respiratory disease (OR 0.65;95% CI, 0.37 to 1.15; p=0.45). Similarly, mortality rate of COVID 19 patients with cancer is close to COVID 19 patients with diabetes (OR 1.32; 95% CI, 0.42 to 4.11; p=0.39), hypertension (OR 1.27; 95% CI, 0.35 to 4.62; p=0.13), and respiratory diseases (OR 0.79; 95% CI, 0.47 to 1.33; p=0.45). Nevertheless, it is worth noting that COVID19 patients with cancer showed higher mortality rate than cardiovascular diseases (OR 0.49; 95% CI, 0.34 to 0.71; p=0.39) (eFigure 3).

#### Quality assessment and publication bias

The quality of the included literature was satisfactory. Insufficient number of patients and incomplete coverage of the population were the main cause of low quality literature (eFigure 1), which we excluded. We performed sensitivity analysis of the included studies to assess their stability. After each study was excluded in turn, we found no significant difference in the results of the analysis (eTable 4,5). The funnel plot and Egger test results showed the risk of publication bias is low (p-value = 0.35) (eFigure 5).

The heterogeneity among our study subgroups was high. In the subgroup mortality analysis, the heterogeneity parameter I^2^ were >50% in all subgroups except group IA (sample size ≤100). So, the random-effects model was deployed.

## Discussion

A comprehensive analysis of cancer prevalence and severe events among patients with COVID-19 can serve as an important reference for oncology clinicians and healthcare policy makers. A few existing studies focused mainly on the prevalence analysis of cancer patients with COVID-19, lacking a comparison of the prevalence or risk for severe events among different areas. ^11,12^We performed a systematic review on the prevalence of cancer and risks of severe events among patients with COVID-19 using a collection of sparse binomial data from published studies. To our knowledge, this is the largest and most comprehensive meta-analysis of the prevalence and fatality analysis of cancer among patients with COVID-19 to date.

In this study, we found that the total prevalence of cancer in COVID-19 patients was 3.97% (95% CI, 3.08% to 5.12%), which was significantly higher than that of the total cancer rate (0.29%) in China. ^47^In other words, approximately 3 in 100 patients with COVID-19 had suffered from superimposed malignancy. The higher prevalence of cancer in COVID19 patients could be due to multiple factors. Firstly, caner is more common among senior patients, who are also susceptible to COVID19. Secondly, due to primary disease and the anticancer treatment, cancer patients are immune compromised, hence they are more vulnerable to viral infection. Thirdly, COVID19 infection has been shown to elevate the serum level of some cancer biomarkers such as CEA and CA19-9 etc., which may increase the cancer detection rate in this population.^10^

The cancer prevalence in COVID19 patients in the city of Wuhan was significantly higher than the rest of China. Meanwhile, it was significantly higher in European and American countries than in Asian countries. The geographical differences cannot be explained by factors like race or climate. It seems to be associated with overall incidence of COVID19 infection in those affected areas. Therefore, effective control of COVID19 infection, especially non-pharmaceutical interventions, could potentially reduce the cancer prevalence.^48^

In addition to the high prevalence, we found that nearly a third of cancer patients with COVID-19 experienced aggravated conditions and were admitted to ICU, which was apparently higher than total COVID-19 patients. Moreover, fatality in cancer patients with COVID-19 was about 24.32% (95% CI, 13.95% to 38.91%), also significantly higher than the rate of total patients with COVID-19 (about 3-7%).^49^ Factors that affected cancer prevalence mentioned above might also explain the worse prognosis of cancer patients with COVID19. Firstly, individuals with cancer are generally older,^50^ which has been shown to express higher levels of angiotensin-converting enzyme II (ACE2), a receptor for SARS-CoV-2 entry into the cell.^51^ Secondly, cancer patients often suffer from immunosuppression, like lymphopenia, which often results in the impairment of the required immune response and may lead to viral propagation, tissue destruction, and progression to severe stages of the disease in ACE2-rich tissues.^52^ Meanwhile, the overload in the hospitals may prevent timely and sufficient medical care for COVID19 patients with cancer, also leading to high fatality. Severe events are not only a major public health issue, but also a serious economic concern. Although health care personnel and resources were heavily allocated to ICU and treating severe cases, the outcomes were far from satisfactory. Thus, prevention of CoVID19 infection using strict quarantine and social distance measures is essential to protect cancer patients from worsening their health conditions.

The outcome of cancer patients with COVID19 was also compared with other morbidities, especially chronic diseases, which were thought as risk factors for severe event in patients with COVID-19. A meta-analysis by Yang et al showed that COVID-19 patients with hypertension, cardiovascular diseases and respiratory disease have higher risk for developing severe complications.^53^ Du et al found that 61.9% patients with hypertension and 57.1% patients with cardiovascular or cerebrovascular diseases developed severe events.^54^ Kang et al found that superimposed heart disease is the highest cause of death among all complications for COVID-19 patients. ^55^From our analyses, the ICU admission rate for cancer patients with COVID19 was comparable with diabetes, hypertension, cardiovascular diseases and respiratory diseases. Meanwhile, the fatality of cancer patients with COVID19 was also similar with other chronic diseases above expect cardiovascular diseases. COVID19 patients with cardiovascular diseases showed higher mortality rate than cancer. Thus, cancer should become one of the notable superimposed comorbidities together with diabetes, cardiovascular and chronic respiratory diseases, contributing to COVID-19 infection and severe events.

Our findings are of values to both cancer patients and oncology clinicians in COVID-19 outbreak countries and territories. Future studies should rely on more detailed analyses. For example, the prevalence and risks of severe events can vary with different cancer types. A recent cohort study by Dai et al. indicated that COVID-19 patients in Wuhan with hematologic cancer had the highest frequency of severe events, which was followed by lung cancer, and patients with metastatic cancer hade much higher incidence of severe events than those with non-metastatic cancers. Moreover, cancer patients who had received surgery tended to have higher risks of severe events than other treatments except immunotherapy. ^56^ We anticipate further investigations on these issues in broader geographic and clinical settings.

### Limitations

Our meta-analysis studied the cancer prevalence and risks of severe events in COVID-19 patients by accommodating fully reported and censored data from diverse literature. But this meta-analysis has limitations. Small-study effects were observed when studies with smaller sample sizes had different incidences and wider CIs for fatality in patients with COVID-19 and cancer. Although the forest plots showed no asymmetry favoring cancer fatality studies, this meta-analysis is subject to publication bias given that all of our analyses were based on publications. In addition, this study is subject to any biases or errors of the original investigators, and the results are generalizable only to patient groups with COVID-19. Meanwhile, it currently cannot summarize the prevalence or risk of severe events of different types of cancer in COVID-19 patients due to the lack of enough data.

## Conclusions

In summary, using the largest dataset to date, our meta-analysis has clearly demonstrated that (1) there is higher prevalence of COVID19 infection in cancer patients, (2) when COVID19 infection occur in a patient with cancer, the patient is more likely to develop severe clinical events. Patients with chronic health conditions require more stringent preventive measures and more delicate therapeutic strategies during the COVID19 pandemic.

## Data Availability

The data used to support the findings of this study are available from the corresponding author upon request.

https://figshare.com/s/8d089dc9c69f2dc483f5

## Author contributions

QS, JXH and EC. Z had access to all the data included in the study and are responsible for the completeness of the data and the accuracy of our analysis. JXH, HSL and ZZ helped to design the study. QS, DYW, and BWC contributed to the statistical analysis and the revision of this manuscript. ZHG and CGZ approved the final manuscript.

## Acknowledgments

This work was supported by Beijing Natural Science Foundation Program and Scientific Research Key Program of Beijing Municipal Commission of Education (KZ202010025047).

